# The impact of Coronavirus disease 2019 (COVID-19) on health systems and household resources in Africa and South Asia

**DOI:** 10.1101/2020.05.06.20092734

**Authors:** Nicholas Davies, Sedona Sweeney, Sergio Torres-Rueda, Fiammetta Bozzani, Nichola Kitson, Edwine Barasa, Simon R Procter, Matthew Quaife, CMMID COVID-19 Working Group, Rosalind M Eggo, Anna Vassall, Mark Jit

**Author notes:** equal contribution (Nicholas Davies, Sedona Sweeney). equal contribution (Rosalind M Eggo, Anna Vassall, Mark Jit). The following authors were part of the Centre for Mathematical Modelling of Infectious Disease COVID-19 Working Group. Each contributed in processing, cleaning and interpretation of data, interpreted findings, contributed to the manuscript, and approved the work for publication: Damien C Tully, Sebastian Funk, James D Munday, Petra Klepac, Rein M G J Houben, Billy J Quilty, Alicia Rosello, Stefan Flasche, Katherine E. Atkins, Simon R Procter, Megan Auzenbergs, Christopher I Jarvis, Nikos I Bosse, Stéphane Hué, Adam J Kucharski, Kathleen O’Reilly, Akira Endo, David Simons, Fiona Yueqian Sun, Graham Medley, Hamish P Gibbs, Rachel Lowe, Kevin van Zandvoort, C Julian Villabona-Arenas, Yang Liu, Sophie R Meakin, Jon C Emery, Arminder K Deol, Eleanor M Rees, Gwenan M Knight, Joel Hellewell, W John Edmunds, Charlie Diamond, Samuel Clifford, Anna M Foss, Amy Gimma, Emily S Nightingale, Carl A B Pearson, Matthew Quaife, Kiesha Prem, Timothy W Russell, Quentin J Leclerc, Sam Abbott, Thibaut Jombart, Kaja Abbas. Corresponding author: Mark Jit, Faculty of Epidemiology and Population Health, London School of Hygiene & Tropical Medicine, Keppel Street, London WC1E 7HT, United Kingdom.

## Abstract

**Background:** Coronavirus disease 2019 (COVID-19) epidemics strain health systems and households. Health systems in Africa and South Asia may be particularly at risk due to potential high prevalence of risk factors for severe disease, large household sizes and limited healthcare capacity.

**Methods:** We investigated the impact of an unmitigated COVID-19 epidemic on health system resources and costs, and household costs, in Karachi, Delhi, Nairobi, Addis Ababa and Johannesburg. We adapted a dynamic model of SARS-CoV-2 transmission and disease to capture country-specific demography and contact patterns. The epidemiological model was then integrated into an economic framework that captured city-specific health systems and household resource use.

**Findings:** The cities severely lack intensive care beds, healthcare workers and financial resources to meet demand during an unmitigated COVID-19 epidemic. A highly mitigated COVID-19 epidemic, under optimistic assumptions, may avoid overwhelming hospital bed capacity in some cities, but not critical care capacity.

**Interpretation:** Viable mitigation strategies encompassing a mix of responses need to be established to expand healthcare capacity, reduce peak demand for healthcare resources, minimise progression to critical care and shield those at greatest risk of severe disease.

**Funding:** Bill & Melinda Gates Foundation, European Commission, National Institute for Health Research, Department for International Development, Wellcome Trust, Royal Society, Research Councils UK.

**Research in context:** *Evidence before this study:* We conducted a PubMed search on May 5, 2020, with no language restrictions, for studies published since inception, combining the terms (“cost” OR “economic”) AND “covid”. Our search yielded 331 articles, only two of which reported estimates of health system costs of COVID-19. The first study estimated resource use and medical costs for COVID-19 in the United States using a static model of COVID 19. The second study estimated the costs of polymerase chain reaction tests in the United States. We found no studies examining the economic implications of COVID-19 in low- or middle-income settings.

*Added value of this study:* This is the first study to use locally collected data in five cities (Karachi, Delhi, Nairobi, Addis Ababa and Johannesburg) to project the healthcare resource and health economic implications of an unmitigated COVID-19 epidemic. Besides the use of local data, our study moves beyond existing work to (i) consider the capacity of health systems in key cities to cope with this demand, (ii) consider healthcare staff resources needed, since these fall short of demand by greater margins than hospital beds, and (iii) consider economic costs to health services and households.

*Implications of all the evidence:* Demand for ICU beds and healthcare workers will exceed current capacity by orders of magnitude, but the capacity gap for general hospital beds is narrower. With optimistic assumptions about disease severity, the gap between demand and capacity for general hospital beds can be closed in some, but not all the cities. Efforts to bridge the economic burden of disease to households are needed.

## Introduction

Coronavirus disease 2019 (COVID-19) is an acute respiratory illness caused by severe acute respiratory syndrome coronavirus 2 (SARS-CoV-2). It emerged in China in late 2019 and spread rapidly before being declared a pandemic by the World Health Organization on 11 March 2020. The first reported cases in South Asia and Africa occurred, respectively, in Nepal on 24 January 2020 and Egypt on 14 February 2020. As of 4 May 2020, cases have been reported in all countries in both regions except for Lesotho^1^.

Mathematical models have projected the likely health burden of a COVID-19 epidemic in many parts of the world^2–4^. These models suggest that an unmitigated epidemic will almost certainly exceed the healthcare capacity of any country. For example, an unmitigated COVID-19 epidemic in the United Kingdom is projected to peak within 11 weeks of the first case and eventually infect 85% of the population, with about 17% of cases occurring in a single week^5^. Some of these infections require hospital treatment (including supplemental oxygen) and a smaller proportion require critical care (with most critical cases requiring mechanical ventilation). Both proportions rise steeply with age and the presence of chronic medical conditions^6^. In the absence of vaccines or effective therapies to reduce the peak incidence of severe COVID-19 outcomes, many countries have resorted to non-pharmaceutical interventions such as travel restrictions and physical distancing measures to delay the spread of SARS-CoV-2, reduce peak incidence and hence avoid overwhelming health systems resources.

However, the potential benefit of such interventions has been questioned in settings beyond North America, Europe and East Asia. Mitigation measures, such as restrictions on economic and educational activities, can have a larger detrimental effect on national and household economies in Africa and South Asia compared to high-income countries. In Africa and South Asia, households are less able to absorb financial shocks due to lower levels of household savings, while governments may not be able to fund compensatory schemes for people losing income due to lack of fiscal surpluses and access to affordable debt. Health system resources are more constrained due to lack of funding. The severity of a COVID-19 epidemic may be exacerbated in these settings by high prevalence of HIV and tuberculosis, poor living conditions, large household sizes and limited hospital capacity, although this may be mitigated by a younger population and lower prevalence of some comorbidities^7^ compared to high-income settings. Hence patterns of healthcare demand are likely to differ from high-income settings, but even a highly mitigated epidemic may exceed healthcare capacity. Projections of the health systems implications of COVID-19 epidemics in such settings are vital for health systems preparedness and choice of mitigation measures.

To date, no study has looked at the COVID-19-related health resource demand and capacity in Africa and South Asia. To address this gap, we used a combined epidemiological-health systems model to understand the spread of SARS-CoV-2 and its impact on hospital and critical care beds, healthcare worker time and costs in Karachi, Delhi, Nairobi, Addis Ababa and Johannesburg. These five megacities were chosen to be all in resource constrained settings, but vary in terms of population demography, healthcare capacity, income distribution and quality of healthcare data.

## Methods

### Infection transmission and natural history

We adapted a previously-described^5^ age-structured stochastic compartmental mathematical model of SARS-CoV-2 transmission (see Figure S1.1 for model flow diagram, and Table S1.1-1.2 for model parameters with references). The population is stratified into 5-year age groups from 0-4 years to 70-74 years, and one age group for 75+ years. The total population size and its age distribution for 2020 are retrieved from WorldPop^8^.

Mixing between individuals is age-stratified, with the age-specific contact rates for each city adopted from synthetic contact matrices constructed for 152 countries^9^. The entire population starts susceptible. To initiate the epidemic, we assume that one infectious person enters the population at time 0. We run 100 simulations for each city, drawing a different value for *R*_0_ for each simulation. Upon contact with an infectious person, a susceptible individual enters the exposed state with a probability depending on the susceptibility of that individual. The susceptibility is determined by the value of *R*_0_, assumed for a given simulation of the model. We assume that *R*_0_ is drawn from a normal distribution with mean 2.68 and standard deviation 0.57, in keeping with values for *R*_0_ measured in settings with no substantial control measures in place^5^.

After spending time in that state, individuals enter either a clinically or subclinically infected state. Clinically infected individuals are those who eventually show clinical symptoms, while subclinically infected individuals are those who show mild symptoms or no symptoms (the latter sometimes referred to as “asymptomatic” individuals). The probability of showing clinical symptoms is age-dependent and estimated from case data from China, Japan, Singapore, South Korea, Italy, and Canada^10^. Individuals in the preclinical state enter the clinical state after several days, and then the removed state. Individuals in the subclinical state enter the removed state directly. The removed state represents all those who have recovered, died, or have been isolated from the community, and hence are no longer infectious. We assume that preclinical infections are as infectious as clinical infections, while subclinical infections are *f* = 0.5 times as infectious as clinical infections, consistent with empirical measurements showing reduced infectiousness of asymptomatic SARS-CoV-2 infections^10–12^.

### Disease severity

There are no published data about the risk of hospitalisation, ICU admission or death following COVID-19 infection in Africa or South Asia. Hence for our base case scenario, we used risks based on our previous work using data from China^5^, which are similar to data from the USA^13^ and Italy^14^ (see Table S2).

However, our admissions data assume that countries strictly follow WHO best practice guidelines for clinical management. In the presence of an extreme surge in healthcare demand, we expect that countries will go into “surge” mode by reducing the proportion of patients who are triaged to admission, and discharging them as soon as possible. Hence we also examined a hypothetical scenario where both patient admission rates and length of stay were decreased by 50% (i.e. 7.3 days in the general ward for severe cases; 3 days in the general ward followed by 4.8 days in ICU for critical cases).

Another complication is that the prevalence of underlying clinical conditions associated with more severe COVID-19 disease (such as chronic kidney disease, chronic heart disease and diabetes) is about half as prevalent in Africa compared to Europe^7^, so severity may actually be lower in the settings we examine. On the other hand, the prevalence of HIV, TB and malnutrition combined are about an order of magnitude greater^7^, and life expectancy in Africa is less than Europe even after adjusting for greater childhood mortality. Hence severity may be greater in these settings. To take into account both possibilities, we varied the odds of hospitalisation (given being a case) and death (given hospitalisation) from 50% to 150% of the base case scenario.

### Health systems and household resources

We estimate the impact of COVID-19 on health system resources and costs incurred by health systems and households. Full details of data, modelling and references used to estimate resources and costs are in Supplemental Material S2

Impact includes basic emergency response management and communication, case reporting, diagnosis and clinical management. These estimates represent an ideal scenario, with services provided according to international guidelines^15,16^, but account for the real costs of providing these services currently in each setting. Hence, we also provide the “surge” scenario above to describe how healthcare systems in these settings may react when faced with overwhelming demand. The costing model assumes no economies of scale at the site level.

For above-service costs, such as training in case management and case reporting, quantities of staff and building/equipment per day were estimated. Service-level costs for diagnosis and clinical management services were estimated using a bottom-up ingredients approach. Unit costs of outputs, such as bed-days and outpatient visits were sourced from a range of primary published and unpublished data, representing the best available estimates of unit costs in each country.

Unit cost estimates represent the economic cost of all resources required to produce health services, including staff time, capital and equipment, drugs, supplies, and overhead costs. These unit costs were combined with normative guidelines and published evidence on quantities of health services required for COVID-19 cases. For all clinical management services, the quantity of bed-days per COVID symptomatic case were defined using available literature on length of stay for severe and critical COVID-19 cases. The number of visits, regiments, supplies, and tests for diagnosis and clinical management services were defined using standard case management guidelines from the WHO^15^ and United Kingdom^16^. Costs of critical case management also included the costs of co-morbidities and complications, the frequency of which were estimated using evidence from China. Additional resources were added to account for costs specific to COVID-19 treatment and infection control, including mechanical ventilation of patients and personal protective equipment for health care workers. Quantities of ventilation and PPE were sourced from WHO standard case management guidelines. We also include a cost of managing death, assuming one body bag per COVID-related death.

We compare resource use against three different capacity constraints: ICU bed-days, general bed-days and health care workers, sourced from WHO global databases and country-specific data sources. Where city specific data were not available, we assume that cities have twice the national average number of hospital beds per capita.

We estimate household costs of illness using an adaptation of a previously published model on the costs of illness associated with symptoms of tuberculosis (persistent cough and fever) from South Africa^17^. We adjusted the model to reflect COVID-19 age distribution based on outputs of the epidemiological model, and used the best-available estimates of household-incurred costs per hospital bed-day for each country. Costs to households included direct medical (ie. user charges, prescription fees) and non-medical (ie. transport, accommodation) out-of-pocket payments, as well as lost income due to sickness. Cases in hospital were also assumed to incur productivity costs for the duration of hospitalization for one caregiver. We also include a household cost of death, consisting of funeral costs and one year of lost income if the deceased was an income-earner. All costs were converted and inflated to 2019 US dollars using GDP deflators and exchange rates from the World Bank. Where prices were adjusted across settings, we used GDP at purchasing power parity (PPP) for non-tradable goods.

## Results

### Cases and deaths

Figure 1 (top row) shows the projected number of cases and deaths due to COVID-19 over the course of an unmitigated epidemic in each city. Supplemental material S3 shows numbers that are projected to occur over time. In each city, cases are predicted to peak around the same time (median 103-119 days after introduction of the first case, depending on city) in an unmitigated epidemic, with hospital and ICU bed demand peaking soon after that (median 111-129 days and 115-132 days respectively). The peak may be reached sooner than three months after the first detected case if surveillance systems are not sensitive enough to pick up the first few sporadic cases.

**Figure 1.**
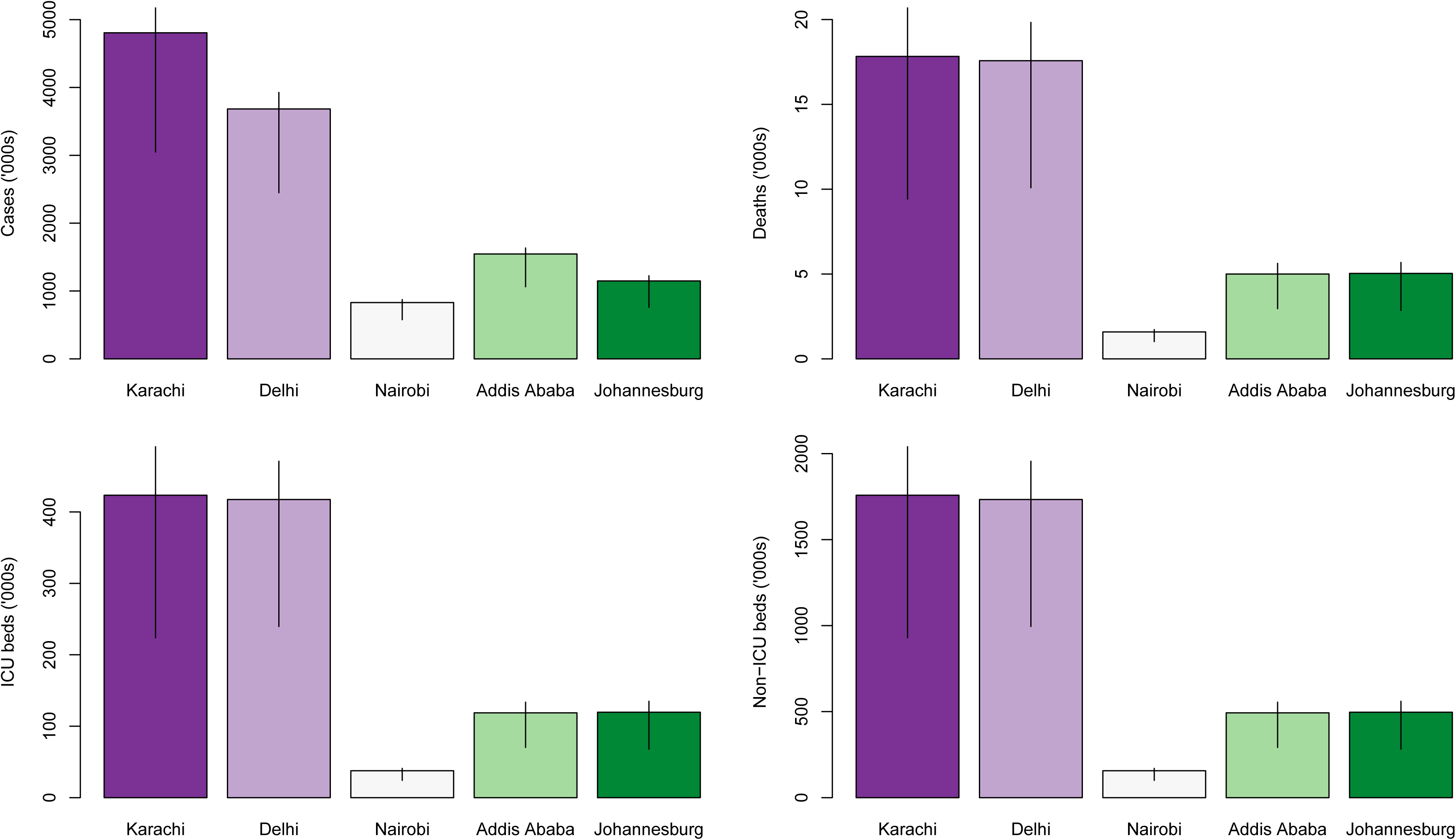
Total projected number of cases, deaths, hospital bed days and ICU bed days over the course of the unmitigated epidemic.

### Health systems resources

Figure 1 (bottom row) shows the number of general hospital beds and ICU beds needed due to COVID-19 over the course of an unmitigated epidemic in each city. Supplemental material S3 shows how the number of general hospital beds, ICU beds and health care workers needed to deal with COVID-19 will change over the course of the epidemic. Peak demand is shown in Figure 4.

The greatest capacity gap is in ICU beds, where median demand in all five cities is expected to exceed bed capacity by 25 (Johannesberg) – 5400 (Addis Ababa) times. The capacity gap for health care staff is also large: 13 (Johannesburg) – 56 (Addis Ababa) times. For hospital capacity, the shortfall is the smallest, with the gap ranging from 3.9 (Nairobi) – 19 (Addis Ababa) times.

### Economic costs

Figure 2 shows the economic costs that are incurred due to COVID-19 by both the healthcare system and households; the costs over time are shown in Figure 3. Median total costs are $43, $56, $66, $33 and $248 per person in Karachi, Delhi, Nairobi, Addis Ababa and Johannesburg respectively. The categories with the highest costs are income losses incurred by households due to patients and their caregivers being unable to work. In comparison, current total/domestic health expenditure per capita are $40/$11 in Pakistan, $62/$16 in India, $66/$24 in Kenya, $27/$8 in Ethiopia and $428/$230 in South Africa^18^.

**Figure 2.**
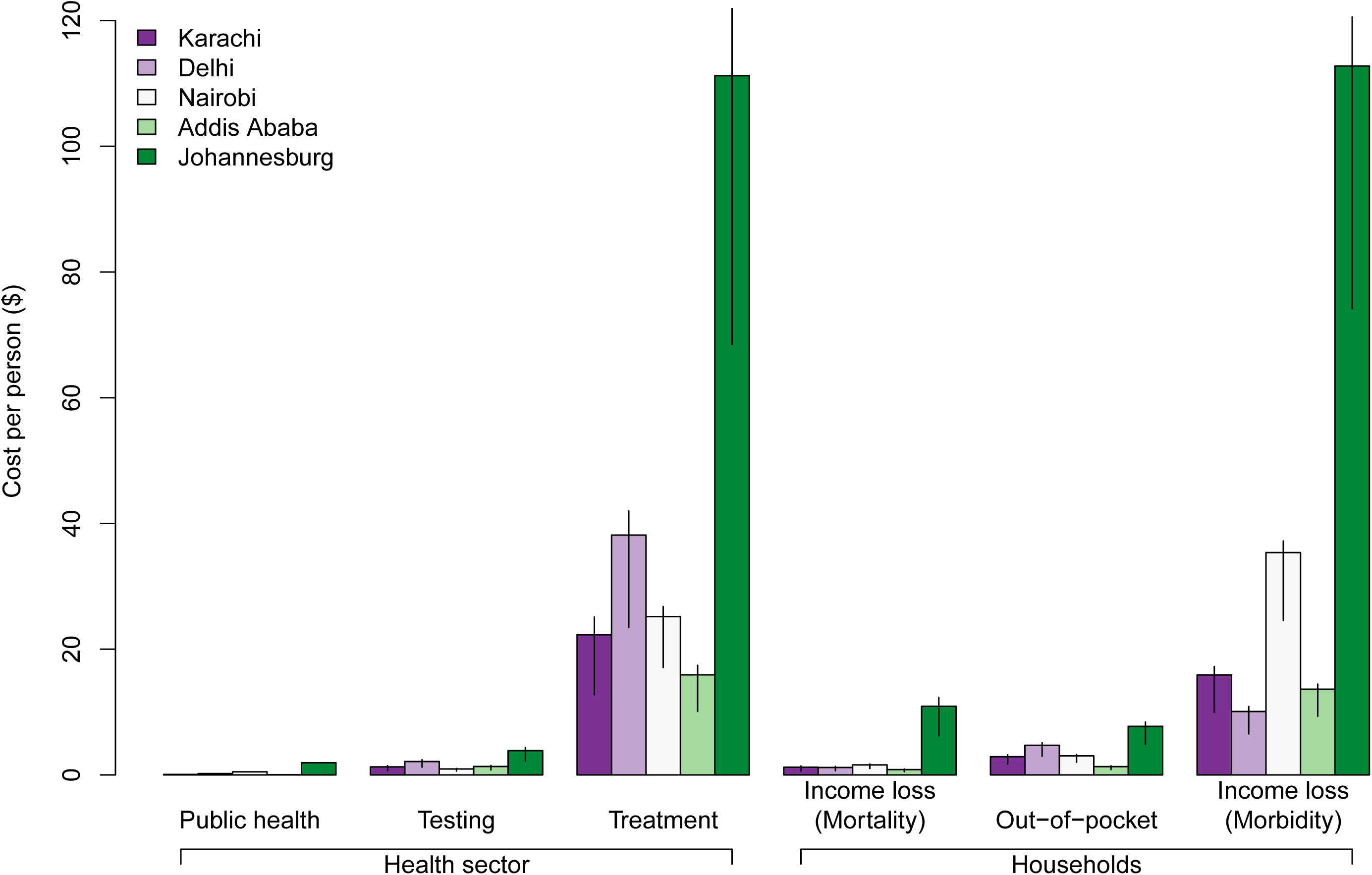
Economic costs per person in the population due to COVID-19 falling on the health system (planning, response, distancing, testing and treatment) and on households (mortality, out-of-pocket, income loss). Error bars show 95% uncertainty range.

**Figure 3.**
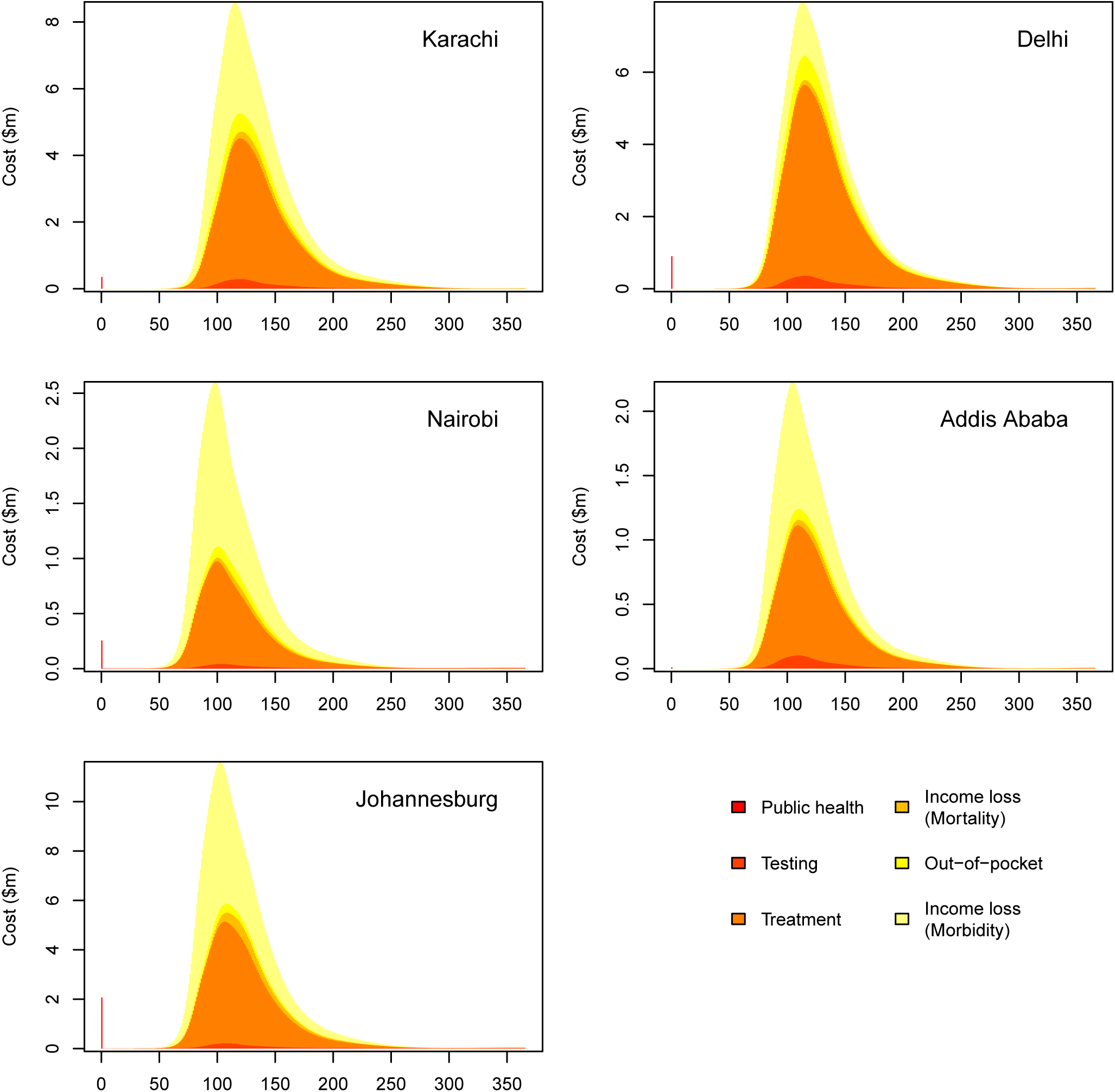
Total economic costs due to COVID-19 by cost category incurred over time.

### Alternative scenarios

Figure 4 shows how peak hospital bed, ICU bed and healthcare worker demand may change if different assumptions about severity and demand per patient are made. Peak demand for ICU beds and healthcare workers remains many times greater than capacity under all scenarios. For general hospital beds, under the most optimistic scenarios, capacity is almost able to meet peak demand in the three African cities.

**Figure 4.**
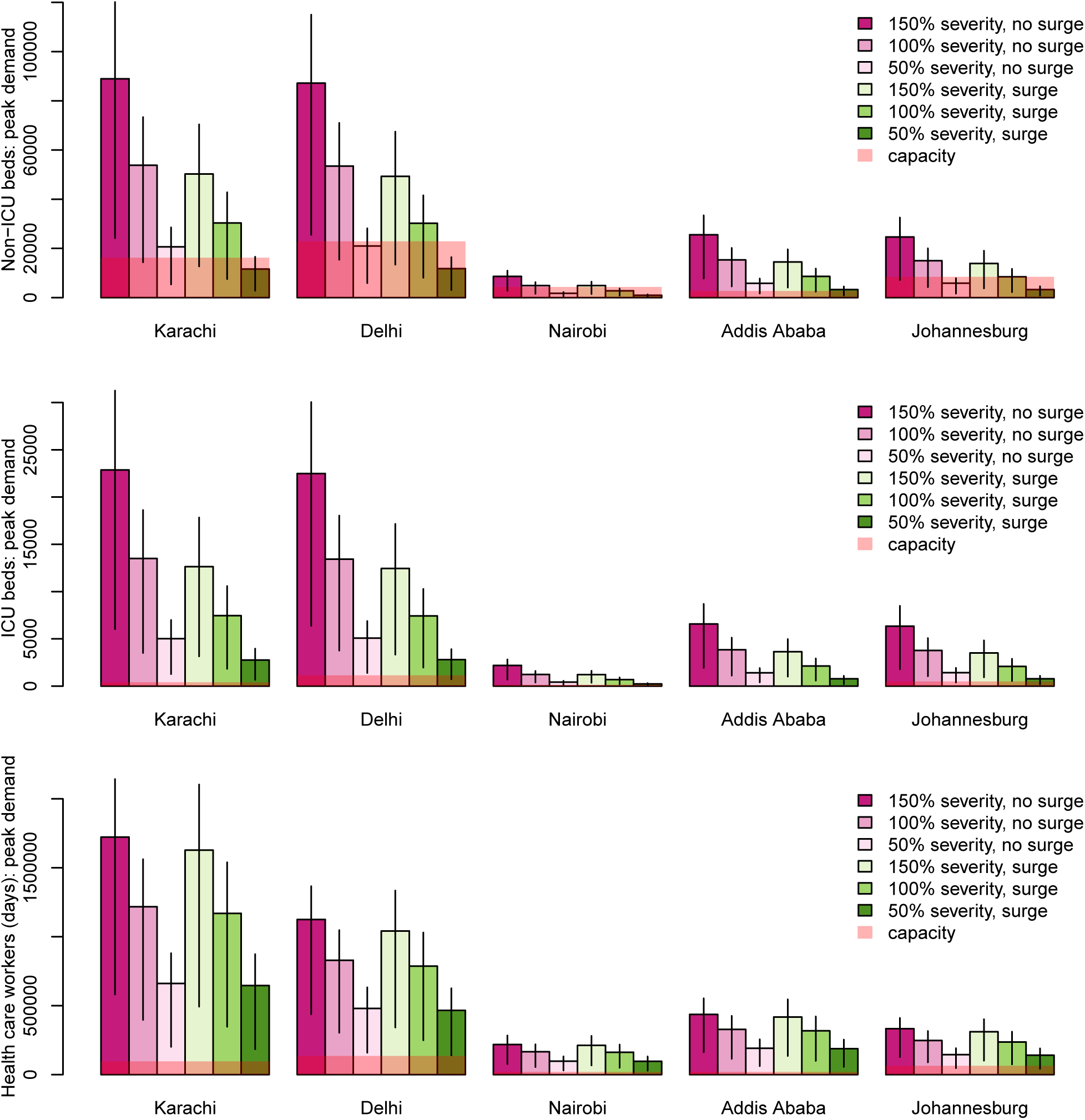
Peak demand for hospital beds, ICU beds and healthcare workers compared to capacity, given different assumptions about severity compared to higher-income settings and bed occupancy (surge assumptions or not). Error bars show 95% uncertainty range.

## Discussion

Using a model that integrates epidemiological, health systems and health economic outcomes, we projected the health service and economic resources needed during an unmitigated COVID-19 epidemic in five cities in Africa and South Asia. We find that demand for ICU beds and healthcare workers will exceed current capacity by orders of magnitude, but the capacity gap for general hospital beds is narrower. Costs falling on both the healthcare sector and on households will also be large, and be close to normal annual healthcare expenditure due to all causes in all the cities except Johannesburg.

With optimistic assumptions about disease severity, the gap between demand and capacity for general hospital beds is closer in some, but not all the cities. If lengths of stay can be reduced and existing beds can be made available for COVID-19 patients, the gap may be small enough to be bridged by a combination of different mitigation measures that flatten the epidemic curve (and hence reduce peak demand for beds). The impact of such interventions on health outcomes in resource-constrained settings has been explored elsewhere^19^. Effective measures include physical distancing, shielding of vulnerable patients (such as older adults with comorbidities) most likely to require hospital admission and construction of field hospitals. Conversely, the shortfall in critical care capacity is likely too large to bridge within the available to prepare for the epidemic peak (and possibly even future waves).

Our modelling takes into account the demographic age profile of the different cities. Hence Nairobi and Addis Ababa are projected to have less severe health care demand shocks compared to the older populations in Johannesburg, Delhi and Karachi more susceptible to severe disease. However, all these cities contain large floating populations such as migrant, informal and casual workers who are not fully in census or even satellite imagery data used by WorldPop to construct city populations^8^. These populations may also bear the brunt of the economic cost of physical distancing and other epidemic mitigation measures. For instance, the lockdown in India (since March 25, extended to May 3) has caused a significant economic shock to millions of migrant and daily wage workers in the informal sector^20^. Hence extremely difficult decisions are needed to trade off the costs of an unmitigated epidemic that we present with the costs of mitigation measures, with both costs falling on both healthcare systems and households.

Several studies have projected the number of cases, hospital admissions and ICU admissions that a COVID-19 epidemic would cause in countries around the world, including Africa and South Asia^34^. Our findings move beyond these studies by (i) considering also the capacity of health systems in key cities to cope with this demand, (ii) considering the healthcare staff resources needed, since these fall short of demand by greater margins than hospital beds, and (iii) considering economic costs to health services and households. These costs include a variety of resources, and hence highlight the inadequacy of simply providing beds, oxygen tanks or ventilators without supporting equipment and staff. For example, costs associated with treatment of severe pneumonia include high-flow oxygen, pulse oximeters and health care staff with training in respiratory medicine in order to effectively recognise and manage hypoxemic COVID-19 patients before they reach a critical condition.

There are a number of important caveats to our findings. The first is that we are examining health care worker capacity considering only doctors, nursing and midwifery staff who are currently available. We do not consider the possibility that staff with less formal training might receive on-the-job training to manage COVID-19 patients. However, we also do not consider that health care workers may themselves be unavailable due to COVID-19 or responsibilities to family members with COVID-19, as has been reported in many countries facing COVID-19 epidemics^21^. Second, our capacity figures show resources available for all healthcare needs, but not all of these can be allocated to COVID-19 patients even if elective procedures are postponed for as long as possible. Indeed, most hospital beds in these cities are near full occupancy, and many health care workers work outside the hospitals. Further work needs to be done to further examine and understand real capacity constraints and the opportunity costs of transferring capacity to COVID-19, where the proportion of hospital admissions that are acute may be higher in high-income settings. Third, our cost results suggest that substantial additional financing is required. However, even where financing may be made available, we do not account for the absorption capacity to use this financing quickly to purchase commodities, construct new infrastructure, train new staff and institute substantial service delivery changes, which will challenge many LMICs, as it has done in high-income countries. Lastly, our economic costs assume that all symptomatic cases would be isolated for the full 7 days recommended by WHO. In practice, this recommendation may not be adhered to by patients with mild disease. Similarly, not all households will necessarily lose income while ill, since some ill individuals may continue to work.

## Conclusion

The consequences of addressing an unmitigated epidemic are substantial for both health sectors and households, risking considerable long-term damage to the economic well-being of both. Addressing COVID-19 in LMICs requires a nuanced and comprehensive approach, which fully considers capacity strengths and constraints in each setting; and trade-offs with broader considerations around poverty alleviation.

## Data Availability

All computer code and model results are available on request.

## Acknowledgments

We thank Allison Portnoy (Harvard) for coding assistance.

## Contributions

MJ, AV and RME conceived the study. SS, ND, STR, FB, NK, EB, SP, SRP and MQ compiled the data sets with direction from AV, MJ and RME. ND, MJ, SS, RME and AV designed the model and conducted the analyses. MJ, AV, ND, SS and RME wrote the manuscript with input from all authors. All authors approved the final version.

## Data sharing

All computer code and model results are available on request.

## References

1 World Health Organization. Coronavirusdisease (COVID-19) Situation Report–105. 2020. https://www.who.int/docs/default-source/coronaviruse/situation-reports/20200504-covid-19-sitrep-105.pdf?sfvrsn=4cdda8af_2 (accessed May 4, 2020).

2 Pearson CAB, Zandvoort K van, Jarvis CI, et al. Projections of COVID-19 epidemics in LMIC countries. 2020. https://cmmid.github.io/topics/covid19/LMIC-projection-reports.html (accessed May 4, 2020).

3 Walker P, Whittaker C, Watson O, et al. Report 12: The global impact of COVID-19 and strategies for mitigation and suppression. 2020. https://spiral.imperial.ac.uk:8443/handle/10044/1/77735 (accessed May 4, 2020).

4 Institute for Health Metrics and Evaluation. COVID-19 Projections. 2020. https://covid19.healthdata.org/ (accessed May 4, 2020).

5 Davies NG, Kucharski AJ, Eggo RM, Gimma A, Group CC-19 W, Edmunds WJ. The effect of non-pharmaceutical interventions on COVID-19 cases, deaths and demand for hospital services in the UK: a modelling study. *medRxiv* 2020;: 2020.04.01.20049908.

6 Verity R, Okell LC, Dorigatti I, et al. Estimates of the severity of coronavirus disease 2019: a model-based analysis. Lancet Infect Dis 2020; 0. DOI:10.1016/S1473-3099(20)30243-7.

7 Clark A, Jit M, Warren-Gash C, et al. How many are at increased risk of severe COVID-19 disease? Rapid global, regional and national estimates for 2020. *medRxiv* 2020;: 2020.04.18.20064774.

8 WorldPop, Center for International Earth Science Information Network (CIESIN). Global High Resolution Population Denominators Project. 2020. https://www.portal.worldpop.org/demographics/ (accessed May 4, 2020).

9 Prem K, Cook AR, Jit M. Projecting social contact matrices in 152 countries using contact surveys and demographic data. PLoS Comput Biol 2017; 13. DOI:10.1371/journal.pcbi.1005697.

10 Davies NG, Klepac P, Liu Y, et al. Age-dependent effects in the transmission and control of COVID-19 epidemics. 2020. https://cmmid.github.io/topics/covidl9/age_hypotheses.html (accessed May 4, 2020).

11 Yi C, Aihong W, Bo Y, et al. The epidemiological characteristics of infection in close contacts of COVID-19 in Ningbo city. Chinese J Epidemiol 2020; 41: E026–E026.

12 Luo L, Liu D, Liao X, et al. Modes of contact and risk of transmission in COVID-19 among close contacts. *medRxiv* 2020;: 2020.03.24.20042606.

13 Bialek S, Boundy E, Bowen V, et al. Severe Outcomes Among Patients with Coronavirus Disease 2019 (COVID-19) — United States, February 12-March 16, 2020. MMWR Morb Mortal Wkly Rep 2020; 69: 343–6.

14 Riccardo F, Ajelli M, Andrianou X, et al. Epidemiological characteristics of COVID-19 cases in Italy and estimates of the reproductive numbers one month into the epidemic. *medRxiv* 2020;: 2020.04.08.20056861.

15 Organization WH. Critical preparedness, readiness and response actions for COVID-19. 2020.

16 BMJ. BMJ Best Practice Coronavirus disease 2019. 2020. https://bestpractice.bmj.corn/topics/en-gb/3000168/pdf/3000168/Coronavirusdisease2019%28COVID-19%29.pdf.

17 Sweeney S, Vassall A, Guinness L, et al. Examining Approaches to Estimate the Prevalence of Catastrophic Costs Due to Tuberculosis from Small-Scale Studies in South Africa. Pharmacoeconomics 2020. DOI:10.1007/s40273-020-00898-3.

18 World Health Organization. Global Health Expenditure Databse. 2020. https://apps.who.int/nha/database (accessed May 4, 2020).

19 Zandvoort K van, Jarvis CI, Pearson C, et al. Response strategies for COVID-19 epidemics in African settings: a mathematical modelling study. *medRxiv* 2020;: 2020.04.27.20081711.

20 Lancet T. India under COVID-19 lockdown. Lancet 2020; 395: 1315.

21 Carding N. Fifth of trust’s medical workforce absent amid covid-19 pandemic | HSJ Local | Health Service Journal. Heal. Serv. J. https://www.hsj.co.uk/workforce/fifth-of-trusts-medical-workforce-absent-amid-covid-19-pandemic/7027329.article (accessed May 5, 2020).

